# DNA methylation in pituitary adenomas: scoping review protocol

**DOI:** 10.1101/2024.04.29.24306412

**Authors:** Morten Winkler Møller, Mathias Just Nortvig, Mikkel Schou Andersen, Frantz Rom Poulsen

**Affiliations:** Department of Neurosurgery, Odense University Hospital, DK-5000 Odense C, Denmark; Clinical Institute, University of Southern Denmark, DK-5000 Odense C, Denmark

**Author notes:** Correspondence to: Morten Winkler Møller –. 2. Registration We intend to file the protocol at medRxiv before conducting the scoping review. 3. Contribution MWM will be the main investigator of the study. The design and search will be performed by MWM. Study inclusion and data extraction will be done by MWM and MJN. Data analysis and writing of the manuscript will be done by MWM. MSA will contribute as an experienced reviewer and if any methodologically challenges his expertise will help resolve these problems. FRP is the most experienced researcher in the group and will be supervising the study. All authors (MWM, MJN, MSA and FRP) will contribute to internal review of the manuscript before submission.

## Abstract

**Objective:** The general objective of this study is to map the current knowledge in DNA methylation in Pituitary Adenomas, and in particular focus on genetic and epigenetic findings and the translation into a clinical setting.

**Introduction:** Pituitary tumorigenesis is currently under major investigation. The current research has turned to epigenomic analysis, investigating whether several epigenetic components like DNA methylation and histone modification can be used as markers for tumorigenesis. Several studies report genes involved in relation to hypo/hypermethylated sites. Others describe differential methylated probes/regions (DMR or DMP) and show subclassification traits. Furthermore, most recent studies aim to cluster samples based on full methylome data (unsupervised cluster analysis), while others chose to assess the most different probes (example 2000 probes) and perform the same data-analysis.

**Inclusion criteria:** In this review, we will include all primary studies on pituitary adenomas analyzed by DNA methylation. No review studies will be included. All papers describing DNA methylation in pituitary adenomas in humans will be included. In addition, all languages will be included. However, both title and abstract needs to be written in English.

**Methods:** Papers will be identified via systematic search using the bibliographic databases: Medline, Embase, Cochrane Library and Scopus. Two reviewers will screen all papers based on title and abstract. All relevant papers will be included for further assessment by the same two reviewers. Full texts from each selected paper will be read, and if relevant regarding the eligibility criteria, papers will be included in the review. Disputes between these reviewers, the paper/papers will be discussed in the entire research-group.

**Funding:** No external funding for this review, funded by in-house resources.

## 6. Rational

Pituitary tumorigenesis is currently under major investigation. Only a couple of mono-genetic mutations have hereditary traits dispositioning for pituitary adenomas (MENIN (1)and AIP (2)). Somatic mutations in GNAS (somatotropic) (3) and USP8 (corticotropic) (4) are the only mutations related to pituitary adenomas. Beyond these two mutations, exom analysis of pituitary adenomas show no other genetic correlation with tumorigenesis (5).

The limited insight provided by genomic analysis have turned the focus into epigenomic analysis (6, 7), investigating whether several epigenetic components like DNA methylation (8) and histone modification (9) can be used as markers for tumorigenesis. Additionally transcriptomic analysis have also aided in the differentiation of subsets of pituitary adenomas (10), but due to the fast technological changes DNA methylation seems to improve the differentiation of subsets while lacking the final step in fully understanding the primary reason for tumorigenesis and the potential clinical impact the current knowledge can provide (11).

This scoping review therefore aims to cover the current knowledge of DNA methylation analysis performed in pituitary adenomas, correlating it with other epigenetic methods, to investigate whether the findings based on DNA methylation are corresponding with other analysis. Furthermore, some publications find general traits regarding subclassifications (12-18), but a general tendency seems to be challenges in coupling these findings with clinical outcomes. Therefore, we perform a scoping review as outcomes seems to be unique regarding each study. Therefore, a systematic review would not be able to conclude any tendencies as outcomes are not comparable.

Several studies report genes involved in relation to hypo/hypermethylated sites. Others describe differential methylated probes/regions (DMR or DMP) and show subclassification traits. Furthermore, most recent studies aim to cluster samples based on full methylome data (unsupervised cluster analysis), while others choose to assess the most different probes (example 2000 probes) and perform the same data-analysis.

Outcomes are reported differently; therefore, the general tendencies and conclusions will be summarized as discrepancies or similarities in interpretation of the results can help illustrate the main acceptance of the role of DNA methylation in a clinical setting.

To our knowledge, the most recent systematic review done on DNA methylation in pituitary adenomas dates to 2013 (19). The present scoping review is therefore highly relevant.

## 7. Objectives

The general objective of this study is to map the current knowledge in DNA methylation in Pituitary Adenomas. This regarding genetic and epigenetic findings and the translation into a clinical setting.

## Methods

### Protocol and registration

Our protocol was structured in line with Preferred Reporting Items for Systematic Reviews for Scoping Review (PRISMA-ScR) (20). The final protocol will be registered on medRxiv before the study is commenced. An a priori search was performed in PROSPERO and medRxiv for pituitary adenomas and DNA methylation, with no protocols found.

## 8. Eligibility criteria

All pre-clinical and clinical primary studies on pituitary adenomas analyzed by DNA methylation in humans will be included. Review studies are excluded. They need to be regarding DNA methylation in pituitary adenomas in humans. Publications describing DNA methylation in pituitary adenomas in any languages will be included, but both title and abstract must be written in English.

## 9. Information sources

We will develop a block search including search blocks for 1. pituitary adenomas and 2. DNA methylation. The search blocks will be developed in Embase (Ovid) and include relevant subject headings and keywords. The search blocks will be translated to the following bibliographic databases: Medline (Ovid), Cochrane Central Register of Controlled Trials (CENTRAL)(Wiley) and Scopus (Elsevier). The searches are reported in the supplementary file. Additionally, forward- and backward-citation searches on included studies will be performed in Scopus (Elsevier).

### Search strategy

The search strategy was developed by MWM and MSA and in collaboration with an information specialist healthcare librarian.

The initial database search strategy was developed for Embase and translated for the other databases (supplementary file – Embase search strategy). Records identified in the database searches will be imported into Covidence (Veritas Health Innovation) and duplicates will be removed. In Covidence, the records will be screened at title/abstract and full text level exported to Endnote.

## 10. Study inclusion

Two reviewers (MWM and MJN) will independently screen all records based on title and abstract. The first 20 records will be screened together to make sure there are consensus regarding the inclusion criteria. Full text versions of relevant publications will be included for further assessment by the same two reviewers. In case of disagreement between reviewers, the paper/papers will be discussed in the entire research-group (MWM, MJN, MSA and FRP).

Any disagreements regarding selection or data will be resolved by discussion in the research group, but if not possible to reach consensus, FRP will have the final word whether a record should be excluded or not.

## 11. Data charting process

A data-charting form in Covidence will jointly be developed by two reviewers (MWM and MJN) to determine which variables to extract. A pilot test of the first five included studies will be performed and the data-charting form will be modified accordingly. Expectantly, data according to DNA methylation; both regarding specific sites (differential methylated positions - DMPs) of hyper- or hypomethylation, in transcription start sites (TSS) or in islands outside genes. Epigenetic modifications will be reported and if specific biologically active region(s) (differential methylated regions – DMRs) is found in the included papers these will be reported. In addition, if any outcomes are reported according to DNA methylation patterns these will be reported in this review. Finally, any prognostic factors or tendencies regarding clinical outcomes corelated with DNA methylation status will be included.

The two reviewers independently chart the data and discuss the results. Any disagreements will be resolved by discussion in the research group, but if not possible to reach consensus, FRP will have the final word on how data should be interpretated.

## 12. Data items

All data for each paper included will be reported as described below:

- Author(s)
- Year of publication
- Origin/country of origin (where the source was published or conducted)
- Aim/purpose
- Population and sample size within the source of evidence (if applicable)
- Methodology/methods
- Intervention type, comparator and details of these (e.g. duration of the intervention) (if applicable). Duration of the intervention (if applicable)
- Outcomes and details of these (e.g. how measured) (if applicable)
- Key findings that relate to the scoping review question/s.

## 13. Synthesis of results

The extent of the empirical research regarding DNA methylation will be displayed by the extracted data from included studies, initially by how the studies was conducted (i.e., retrospective vs. prospective, which platforms was used (illumine or others), handling of the tissue (fresh frozen or formalin fixed, paraffin embedded (FFPE), and other differences between the studies). Description of specific differences in pituitary DNA methylation profiles from CpGs to DMPs to DMRs will by annotated. Specific gene alterations or clinical manifestations according to DNA methylation will be noted as well. Difficulties using DNA methylation or pitfalls regarding analysis of tissue with this method will also be described.

Finally, lack of evidence or the need for further studies regarding DNA methylation will be noted as well since this scoping review not only intend to report the current knowledge but also identify areas in which new research is needed.

## Data Availability

All data produced in the present study are available upon reasonable request to the corresponding author

## 14. Acknowledgements

We thank Andreas Lundh from Cochrane Denmark and Centre for Evidence-Based Medicine Odense for commenting on the protocol. The search strategy was developed in collaboration with an information specialist. We thank professional healthcare librarian Mette Brandt Eriksen, University of Southern Denmark. The funding by Novo Nordisk Fonden is greatly appreciated.

## 15. Conflicts of interest

The other authors report no conflicts of interest.

## Supplementary – Search strategy Embase

**Embase Classic+Embase <1947 to 2024 April 17>**

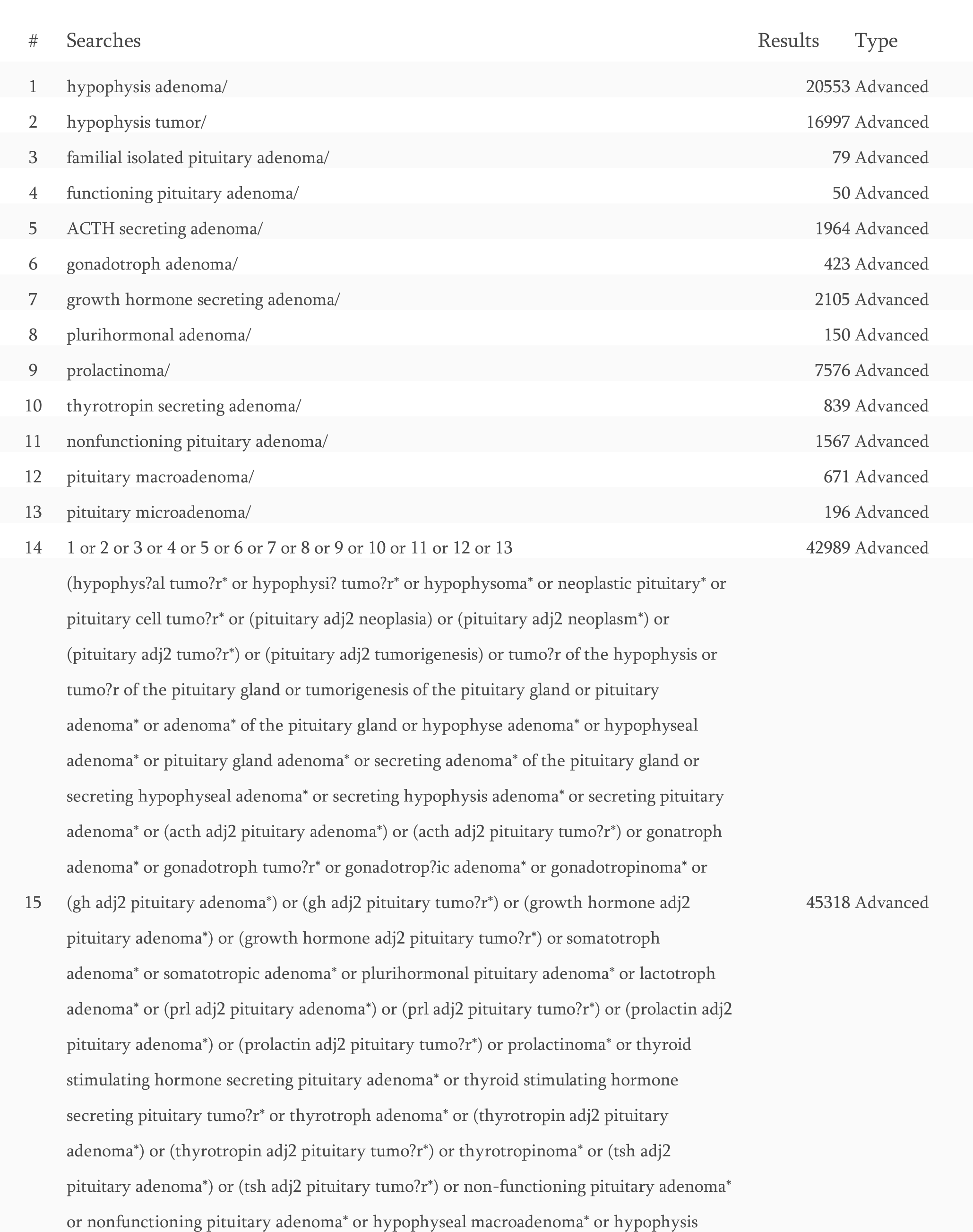
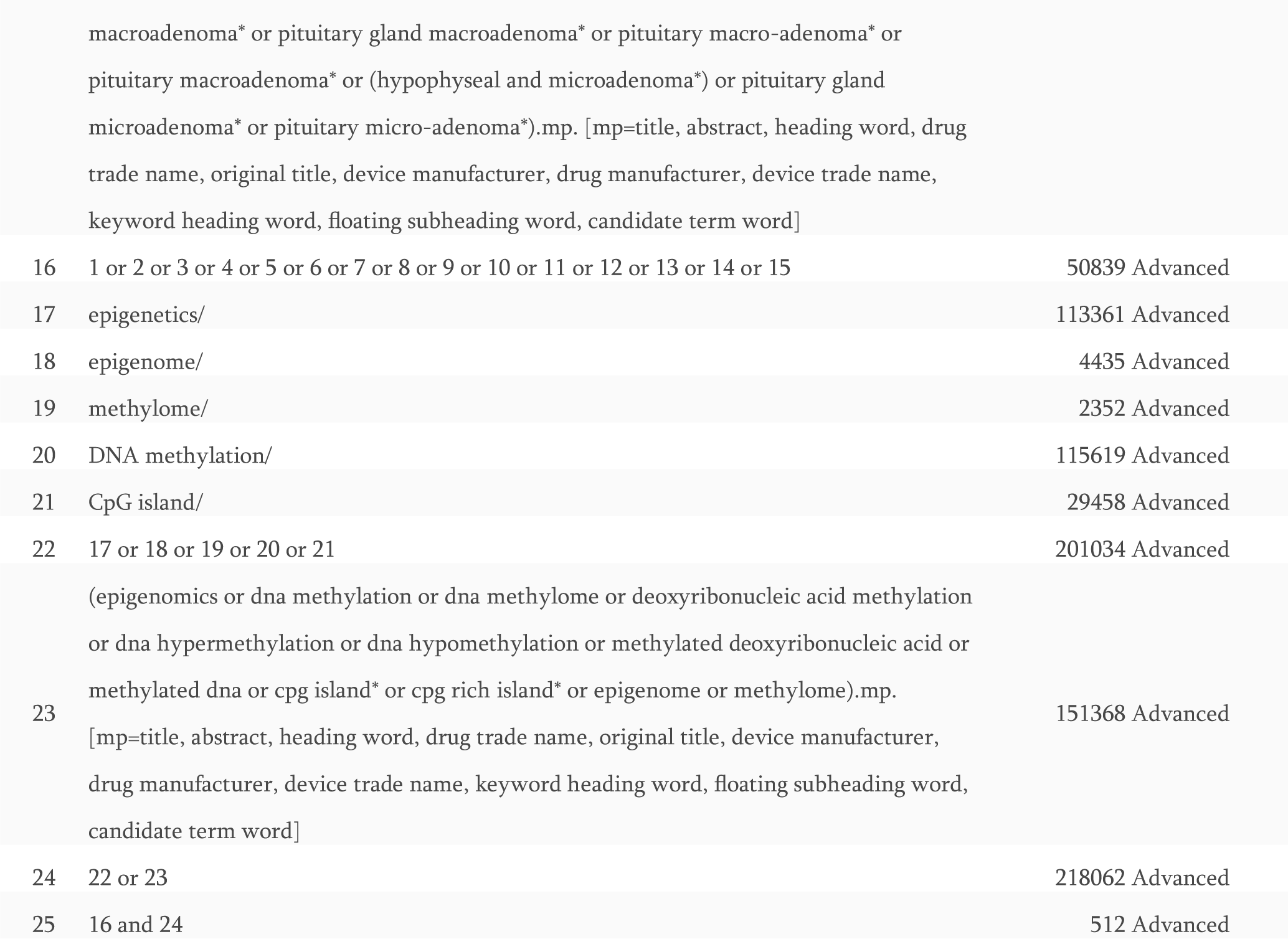

## Notes

### Competing Interest Statement

The authors have declared no competing interest.

### Funding Statement

This study did not receive any funding

